# Measuring effects of ivermectin-treated cattle on malaria vectors in Vietnam: a village-randomized trial

**DOI:** 10.1101/2023.02.09.23285706

**Authors:** Estee Y. Cramer, Nguyen Xuan Quang, Jeffrey C. Hertz, Do Van Nguyen, Huynh Hong Quang, Ian Mendenhall, Andrew A. Lover

**Affiliations:** Department of Biostatistics and Epidemiology, School of Public Health and Health Sciences, University of Massachusetts-Amherst, USA; Institute for Malariology, Parasitology and Entomology, Ministry of Health, Quy Nhon, Vietnam; U.S. Naval Medical Research Unit TWO, Singapore; Programme in Emerging Infectious Diseases, Duke-NUS Medical School, Singapore

**Keywords:** Malaria, mosquitoes, malaria elimination, vector-borne disease control

## Abstract

**Background:** Malaria elimination using current tools has stalled in many areas. Ivermectin (IVM) is a broad-antiparasitic drug and mosquitocide that has been proposed as a tool for reaching malaria elimination. Under laboratory conditions, IVM has been shown to reduce the survival of *Anopheles* populations that have fed on IVM-treated mammals. Treating cattle with IVM has been proposed as an important contribution to malaria vector management, however, the impacts of IVM in this animal health use-case had been untested in field trials in Southeast Asia.

**Methods:** Through a randomized village-based trial, this study aimed to quantify the effect of IVM-treated cattle on anopheline populations in treated vs. untreated villages in Central Vietnam. Local zebu cattle in six rural villages were included in this study. Cattle were treated with IVM at established veterinary dosages in three villages and in three additional villages, cattle were untreated as controls. The mosquito populations in all villages were sampled using cattle-baited traps for six days before, and six days after a 2-day treatment IVM-administration (intervention) period. Vector species were characterized using taxonomic keys. The impact of the intervention was analyzed using a difference-in-differences (DID) approach with generalized estimating equations (Poisson distribution with bootstrapped errors).

**Results:** Across the treated villages, 1,112 of 1,527 censused cows (73% overall; range 67% to 83%) were treated with IVM. In both control and treated villages, there was a 30% to 40% decrease in total anophelines captured in the post-intervention period as compared to the pre-intervention period. In the control villages, there were 1873 captured pre-intervention and 1079 captured during the post-intervention period. In the treated villages, there were 1594 captured pre-intervention, and 1101 captured during the post-intervention period. The DID model analysis comparing total captures between arms was not statistically significant (p = 0.67). Secondary outcomes of vector diversity found that in four villages (two treated and two control) there were statistically significant changes in the anopheline population diversity (p < 0.05) based on Shannon’s diversity index. Two villages (one treated and one control) had a statistically significant increase in diversity and two villages (one treated and one control) had a significant decrease in population diversity (p < 0.05). There were no clear trends in treated or untreated vector population evenness or richness estimates.

**Conclusions:** Unexpectedly large decreases in trapping counts post-intervention across all study villages impacted the ability of this study to quantify any differential impacts. As such, the results of this study do not provide evidence that treating cattle in villages with IVM reduces nightly captures from cattle-baited traps of female anopheles mosquitoes when compared to control villages. The lack of differential impacts may be due to several factors including the short half-life of IVM, crossover in mosquito populations between treated and control villages, feeding preferences of the mosquitoes, and mass-action effects from extensive mosquito trapping. Future studies should plan to treat at least 80% of the cattle in the village and evaluate the relationship between dose-density and mosquito prevalence. Additional studies should investigate whether IVM differentially impacts vector species at a population level.

## Introduction

Major progress has been made in many areas globally toward malaria elimination; through multiple elimination initiatives, malaria incidence in Vietnam has declined substantially in the last two decades; from 2000 to 2019, there has been a 95% reduction in cases and a 96% reduction in deaths caused by malaria.^1^ The achievements in malaria reduction prompted the Government of Vietnam to set the goal of *Plasmodium falciparum* elimination by 2025 and malaria elimination due to all *Plasmodium* species by 2030 in Vietnam.^2^

Globally, malaria control and elimination programs are focused on high coverage of long-lasting insecticide-treated nets (LLINs), indoor residual spraying (IRS) in some areas, universal access to artemisinin-based combination therapy (ACT), and rapid diagnostic tests,^3^ however, these tools may not be sufficient to achieve malaria elimination in all settings. Specifically, progress towards elimination has been stalled due to “residual transmission” which may be driven by outdoor biting vectors, changes in vector bionomics, and decreased sensitivity to insecticides.^4^ These problems are especially complex in the Greater Mekong Subregion, where many vectors have been implicated in transmission, and where peri-domestic vector feeding is common.^5^

One tool proposed to accelerate malaria elimination is ivermectin (IVM). IVM is an inexpensive and non-toxic helminthicide and mosquitocide with a well-established regulatory environment.^6^ While IVM is lethal to invertebrates, it has very limited toxicity in mammals.^7^

Due to IVM’s lipophilic characteristics, IVM undergoes little metabolism and is excreted nearly unchanged after administration in cattle or humans. While this drug is practical for use in animals being raised for consumption, it has a short half-life. Therefore, cattle need repeated doses in order to reduce the survival rates of blood-feeding arthropods including mosquitoes.^8^ Because of the highly desirable characteristics of IVM, several prior studies have tested the impact of using this drug to increase mosquito mortality.^9–13^ All studies published to date show that under laboratory conditions, mosquito populations that have taken blood meals from IVM-treated cattle have major decreases in survival rates, especially those that feed just a few days after the cattle are treated.

Though treating cattle with IVM has been demonstrated to be an effective mosquitocide in laboratory conditions, to our knowledge, no field trials have been published examining the impact of IVM treatment in cattle on a village-level. By focusing on zoonotic feeding by vectors, a targeted program has the potential to reduce the overall peridomestic anopheline populations in suitable contexts. Importantly, in the GMS and in many other endemic areas, livestock are often kept in close proximity to residential areas, providing a clear rationale for this targeted intervention.^14^

The GMS is an ideal setting for this elimination tool because in this region, many anopheline species display zoophilic and anthropophilic behavior.^15^ Anopheline species commonly found in the area include *Anopheles vagus, An. sinensis*, and *An. peditaeniatus*.^16^ All three species are generally exophagic. *Anopheles vagus* is mainly zoophilic in Southeast Asia, while *An. Sinensis* and *An. peditaeniatus* feed on both humans and cattle. *Anopheles peditaeniatus* is largely anthropophilic.^17^ However across the region there is extensive spatial variation in feeding preferences.^18^ Additionally, many cattle are routinely treated for infections such as helminths and heartworm, thus there is already an established system in place to administer medication to these animals.^19^

Using a randomized village-based trial, this study quantifies the effect of IVM-treated cattle on the anopheline populations in treated vs. control villages in rural areas in central Vietnam. We hypothesize that villages with IVM-treated cattle will have a larger decrease in anopheles mosquitoes captured during the post vs. pre-intervention period as compared to control villages without IVM-treated cattle.

## Materials and Methods

### Study Site

This study was conducted in 6 rural villages in Krông Pa District of Gia Lai province in Central Vietnam [Chính Đơn (CD), Hòa Mỹ (HM), Ơi Jit (OJ), Ơi Đăk (OD), H Yú (HY), and H Lang (HL)]. In these semi-rural villages, livestock are generally penned adjacent to, or directly under, raised homes (Figure 1). These specific villages were chosen in close consultation with local health staff, who were familiar with the area’s cattle-rearing practices and had strong links to local village leaders.

**Figure 1:**
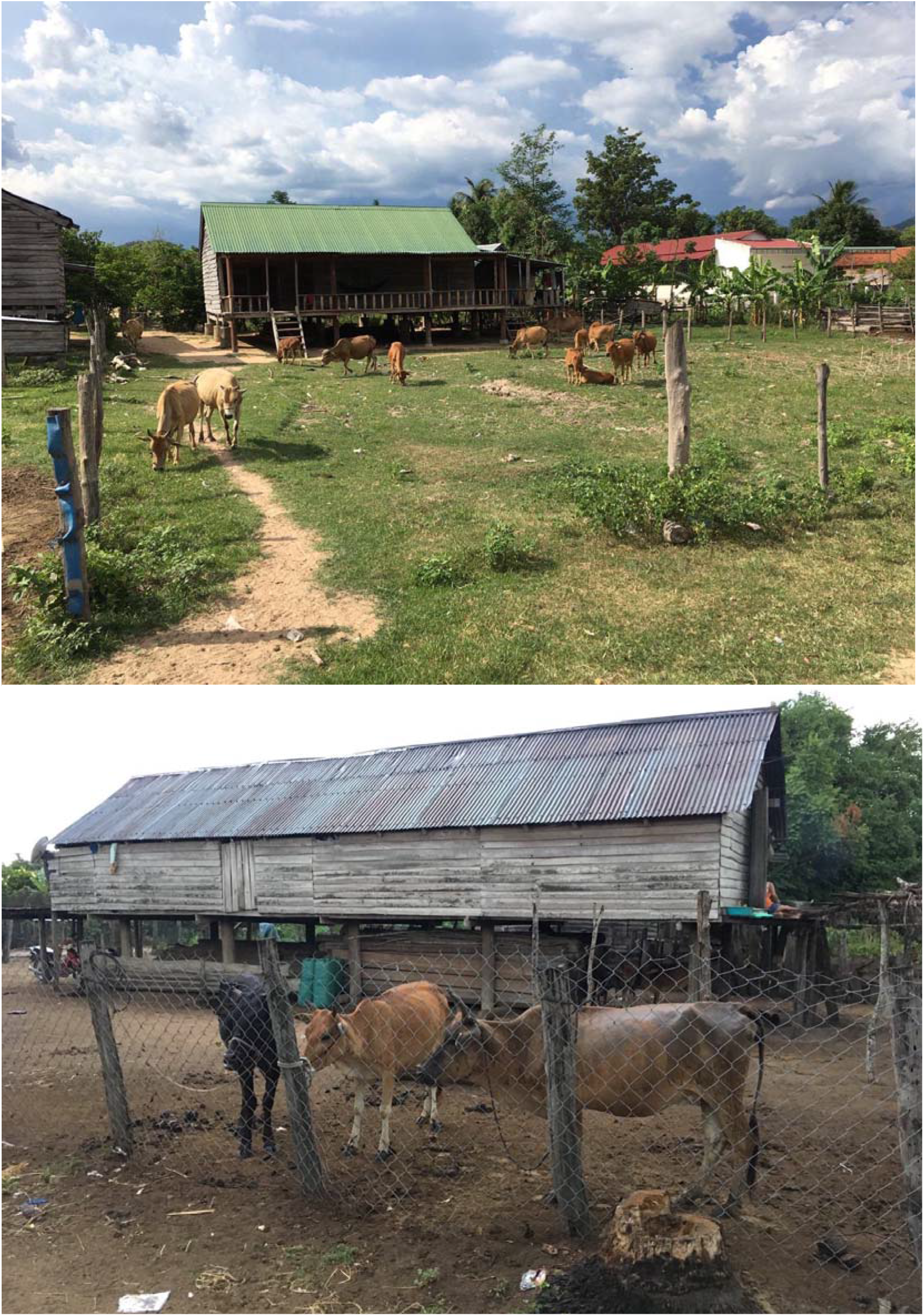
Cattle in close proximity to households, Central Vietnam.

### Study Design

To quantify the relationship between treating cattle with IVM and mosquito prevalence, a village-based randomized controlled trial was implemented. In this trial, three villages (HM, OJ, and HL) had their cattle treated with IVM, and three villages (CD, OD, and HY) served as controls. The villages were chosen for treatment using a random draw (Stata, ralloc).^20^

The study design was constrained by the logistics of trapping cycles and funding available for this pilot study. The number of villages and sampling sites required within each arm for valid statistical comparisons was estimated using a simulation-based approach, as direct analytical methods have not been developed for repeated-measures (longitudinal) experimental designs. Simulations were performed using Stata software (version 15, College Station TX, USA); power calculations for varying numbers of villages, and sample sizes per village are shown in Supplemental Table 1. The sample size for the primary evaluation at the endline survey was determined based upon the power to detect a difference between treated and control villages, assuming a 50% reduction in total nightly *Anopheles* spp. vector catches from cattle-baited traps.

Simulations were parameterized using capture rates from trapping data from the Institute of Malariology, Parasitology, and Entomology (IMPE) (2016; unpublished), published data collected in Lao PDR,^21^ and cattle-baited trapping data from Cambodia.^22^ Prior trapping data from Krông Pa using CDC traps found a mean of 3.5 vectors/night (median 1; SD= 6.5), and data from Cambodia suggest that cattle-baited traps (CBTs) have nightly capture rates that are 10-20-fold higher.^22^ With an assumption of a mean of 35 captures per trap-night, and with a between-village variance of 0.10, to detect a 50% difference in captures with 80% power at a 5% significance level, a total of six trapping nights (three pre-intervention and three post-intervention) are required (Supplemental Table 1).

The primary study outcome was a comparison of pre- and post-intervention trap-night totals of all female anopheline species captured via centralized CBTs (with one per village site). These traps were chosen to maximize the study power, as CBTs have been shown to capture 10- to 20-fold more vectors relative to CDC traps in the GMS.^22^ To determine the impact of IVM treatment on mosquito populations, mosquitoes were captured for a period prior to IVM administration (pre-intervention period). Following baseline mosquito collection, there was a two-day wait period followed by IVM administration to cattle in the three treatment villages over a two-day period. This was followed by another two-day wait period, then another 6 days of mosquito trapping in all 6 villages (post-intervention period). Overall, a total of 72 trapsite-nights of vector trapping were conducted during the study.

### Mosquito capture

To capture anophelines, CBTs^23^ were used from 18:00 to 6:00 the following morning; traps were swept with hand aspirators during collections. For each CBT trap-night, a new cow was randomly selected in each control village. In treated villages, a minimum of four male cows were left untreated, and then each night a new random selection was made from the combined total of the four untreated males plus all other untreated cows (pregnant, young calves, etc.).

### Cattle treatment

Prior to treating the cattle, a short questionnaire was administered to the head of each household to determine the total number of cows that the household owned and the total number of cows that were pregnant, currently ill, or under 1 year old. After owner consent, all cattle eligible for treatment were injected with a standard veterinary dose of 0.2 mg/kg IVM per total body weight of the cattle with a 1% IVM dosage by staff from the local animal health workers using girth-weight charts,^24^ and validation for Asian breeds.^25^

Treatments were conducted across all treated villages over the same two-day period. Veterinary-grade “Vimectin” (Vemedim Corporation; Can Tho, Vietnam) was administered by joint teams from IMPE and local animal health staff. Cattle in control villages were not treated with IVM or a placebo during this study, hence, no blinding was possible.

### Mosquito classification

The collections were identified using a standard key to the mosquitoes of Thailand.^26^ Identified female mosquitoes were placed into individual cryotubes and stored at −20° C or −80° C until processing. Sporozoite rates were not analyzed in this study, as very low indexes are the norm throughout the GMS (1/1000 or 1/2000),^27^ which precludes any valid statistical inferences.

### Statistical Analysis

To evaluate the impact of IVM in the treated vs. control villages, a difference-in-differences (DID) model was used. Specifically, differences in trapping totals were quantified between study arms and interventional periods using generalized estimating equations (GEE), with a Poisson distribution and error-clustering at the village-level. Standard errors for estimates were determined using a bootstrap method with 5000 replicates. The Poisson distribution was used due to over-dispersed trapping counts. Statistical significance of IVM treatment was determined as having a p-value less than 0.05. Main analyses were run using Stata software (version 16, College Station TX, USA) and other analyses were run in R.^28^

### Species diversity calculation

To assess the potential impacts of IVM treatment on anopheline species diversity at the village-level, the commonly used ecological metric of Shannon’s diversity index was used. The minimum value is 0, which occurs when only one species is present, and the maximum value is log(k) where k is the number of species in a population. The maximum value indicates higher species diversity and occurs when there is an equal number of organisms present for each species.^29^ Shannon’s diversity metric is an aggregation of the measures of species richness (a count of the number of species in a population) and species evenness (measured as p_i_ log p_i_) where p_i_ is the relative proportion of a species in a population.

The diversity index for each village was estimated before and after the intervention period, and Hutcheson’s t-test was used to determine if statistically significant differences exist in the species diversity, across the pre- and post-intervention periods.^30^ The Bonferroni correction was used to adjust for multiple testing across the village sets. Additionally, Shannon’s diversity index was calculated for each individual day of mosquito trapping in each village to assess longitudinal changes in each study site. In addition to species diversity, species richness and evenness were also evaluated for each village at each date.

### Ethical clearance and consent

IACUC approvals were obtained from the National University of Singapore (ref: B18-0303), and the University of Massachusetts (2019-0011); and all local regulations were followed. Cattle owners were counseled not to sell meat or dairy for up to 28 days post-intervention. All owners willing to enroll their cattle were informed of these limitations in writing, with follow-up by local ministry of agriculture staff to ensure adherence. Treatment of cattle with subcutaneous injections of ivermectin is routine policy in animal health, and the drug is fully approved in Vietnam for veterinary use (National Guidelines for Animal Health, 2016). No adverse events were reported.

## Results

### Treatment coverage

During the intervention period, all eligible and owner-consented cattle in the three treated villages (HM, OJ, and LA) were injected intravenously with IVM at standard dosing. Over two days, a total of 1,112 cattle were treated with 18,761 units of IVM (1,993 doses in HM, 6,200 in OJ, and 10,568 in HL). Total cattle coverage was over 80% in HM and OJ (81.1% and 83.8% respectively) and 66.8% in HL (Table 1). Using approximate areas determined from satellite imagery (Google Maps), the estimated total village size was determined in m^2^; these areas are shown in Table 1.

**Table 1:** IVM dosages administered, treatment coverage, and density in three villages, Central Vietnam, 2019.

| Treated Village | Total Cattle treated | Total ivermectin dose units dispensed | Estimated cattle coverage (%; 95% CI) |
| --- | --- | --- | --- |
| Hòa Mỹ (HM) | 120 | 1,993 | 81.1<br>(73.8 - 87.0) |
| Ơi Jit (OJ) | 363 | 6,200 | 83.8<br>(80.0 - 87.2) |
| H Lang (LA) | 629 | 10,568 | 66.8<br>(63.7 - 69.8) |

### Mosquitoes captured

A total of 5,647 female anophelines were trapped from September 10^th^ to October 6^th^, 2019, of which, 3,467 were captured before the intervention, and 2,180 were captured post-intervention. These totals correspond to a mean of 96 per trapsite-night pre-intervention, and a mean of 61 per trapsite-night post-intervention; all substantially larger than the minimum required for sufficient study power.

Across all villages, the largest number of mosquitoes captured was in CD during the pre-intervention period; 923 mosquitoes were captured, with a median of 141.0 captured per night. The second largest number captured was in OJ during the pre-intervention period with a total of 919 specimens captured, with a median of 161.5 per night. In five out of six villages, there was a decrease in the total number of female *Anopheles* captured during the post-intervention period compared to the pre-intervention period. The only exception was the control village, HY, which had a total of 263 mosquitoes captured pre-intervention and a total of 340 mosquitoes captured post-intervention (Figure 2). Morphological identification captured 18 unique *Anopheles* species across all sites. HY had 14 species, CD had 13, HM, OD, and OJ had 12 unique species, while HL had 10 unique species.

**Figure 2:**
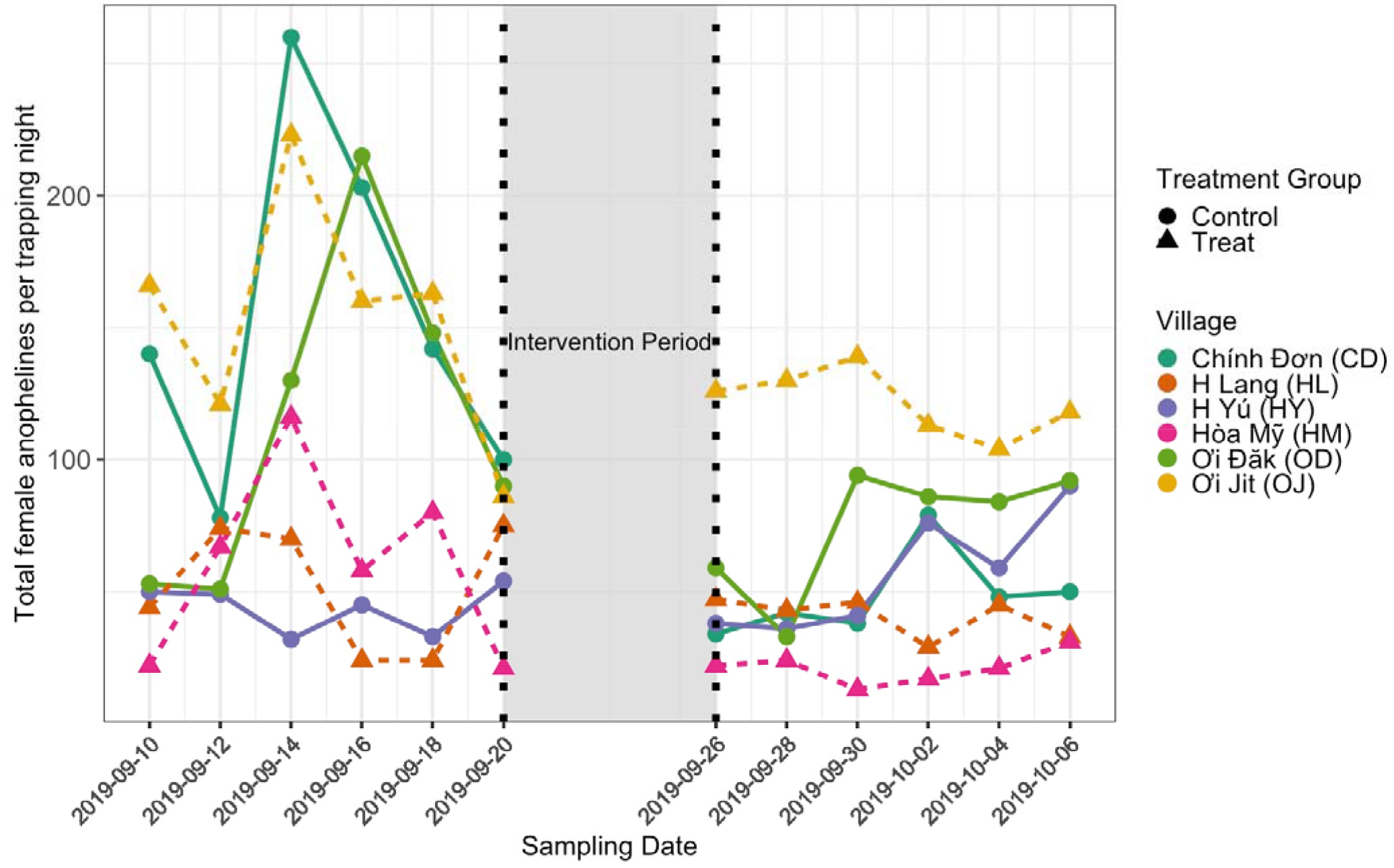
Trial overview, showing *Anopheles* species captured each night before and after intervention in all villages. Each village is represented by a color. Dotted lines and triangles represent villages that received IVM treatment for their cattle, and solid lines with circles represent villages that were not treated with IVM.

The most commonly collected mosquito species across all sites was *An. peditaeniatus* (1,812 captured during the pre-intervention and 1,047 captured post-intervention). The next most commonly captured species were: *An. aconitus* (835 pre- and 522 post-intervention), *An. sinensis* (326 pre and 194 post), and *An. vagus* (242 pre and 113 post) (Figure 3). In CD, HM, OD, and OJ, the number of species captured during the pre- vs post-intervention period decreased (Table 3). In HL the number of different species remained stable across the study, while in HY there were more unique species captured during the post-intervention period.

**Figure 3:**
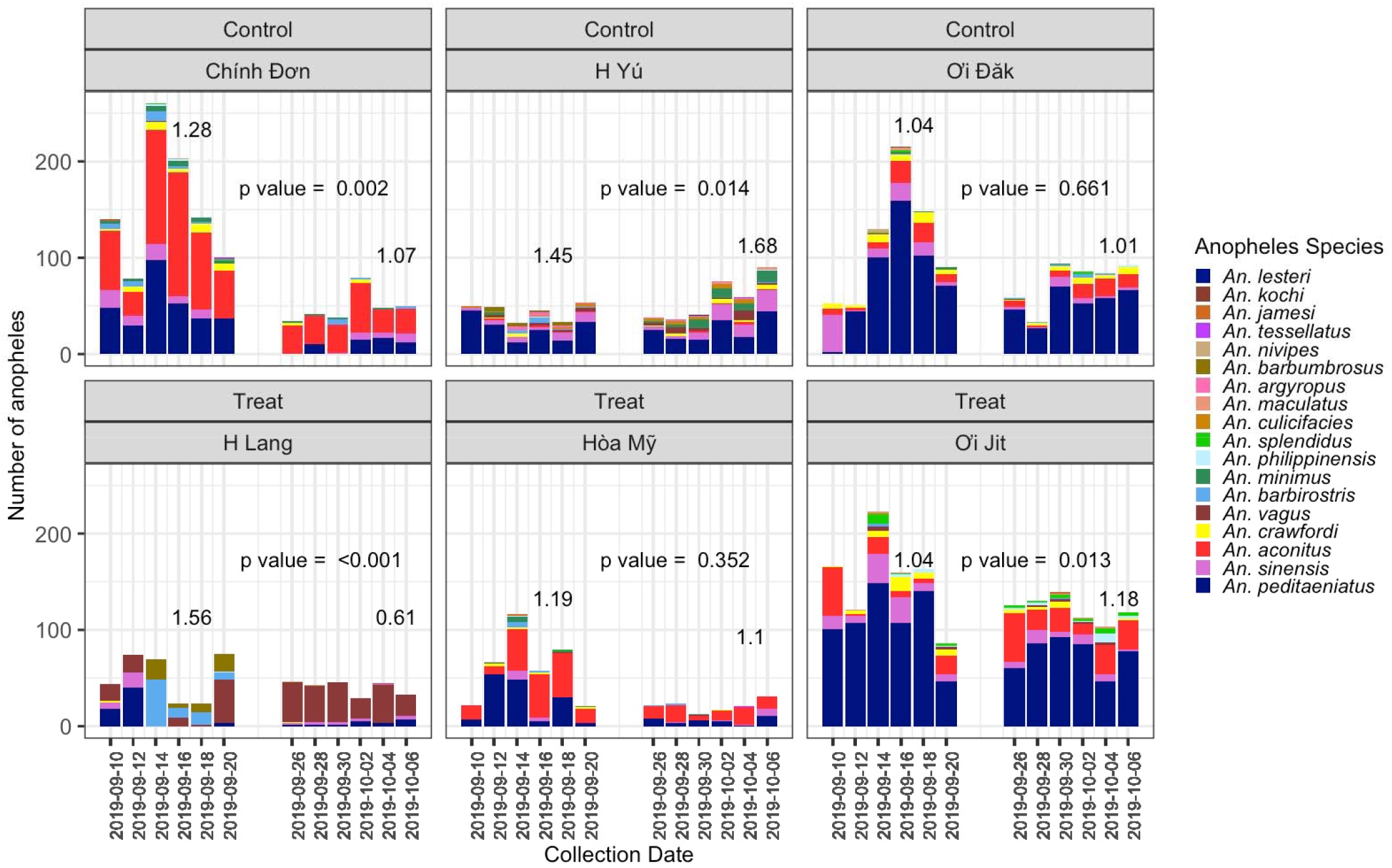
Count of *Anopheles* species captured nightly Count of female *Anopheles* mosquitoes captured with a cattle-baited trap stratified by mosquito species. Numbers above each treatment and control period are the calculated Shannon’s Diversity Index. p values included were calculated with a Hutchinson’s t-test and corrected using a Bonferroni method.

### Impact of treatment on total mosquito captures (primary outcome)

In both the treated and control villages, there was a marked reduction in mosquito captures during the post-intervention period compared to the pre-intervention period. The reduction in the treated villages (30.9%) was less than the reduction in the control villages (42.4%). A statistical model was used to determine whether there was a significant impact of the intervention during this period. Based on model estimates, there was no evidence for a statistically significant difference in sampled mosquito density between the treatment and control groups, with an interaction term (as incidence rate ratio) of 1.19, (p = 0.67) (Table 2).

**Table 2:**
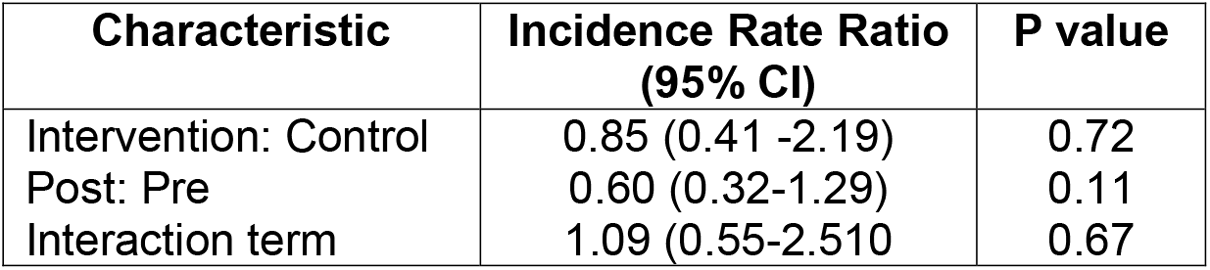
Results from the Difference in Differences GEE model Results from the generalized estimating equation with a Poisson distribution. Confidence intervals and p-values were calculated using a bootstrapped approach. The most relevant data point, the interaction term, is used to show the difference in mosquito populations due to both time and intervention. As shown by the small rate ratio and the insignificant p-value, IVM treatment did not significantly impact the mosquito prevalence in the treated groups compared to the control groups.

### Impact of treatment on Anopheles species diversity (secondary outcomes)

In each village, there were measurable changes in the species diversity during the study period; these changes were significant (p < 0.05) in four villages: CD, HY, LA, and OJ. When evaluating the components of Shannon’s index (species richness and species evenness) there were no clear trends in these metrics after IVM administration (Figure 5).

**Figure 4:**
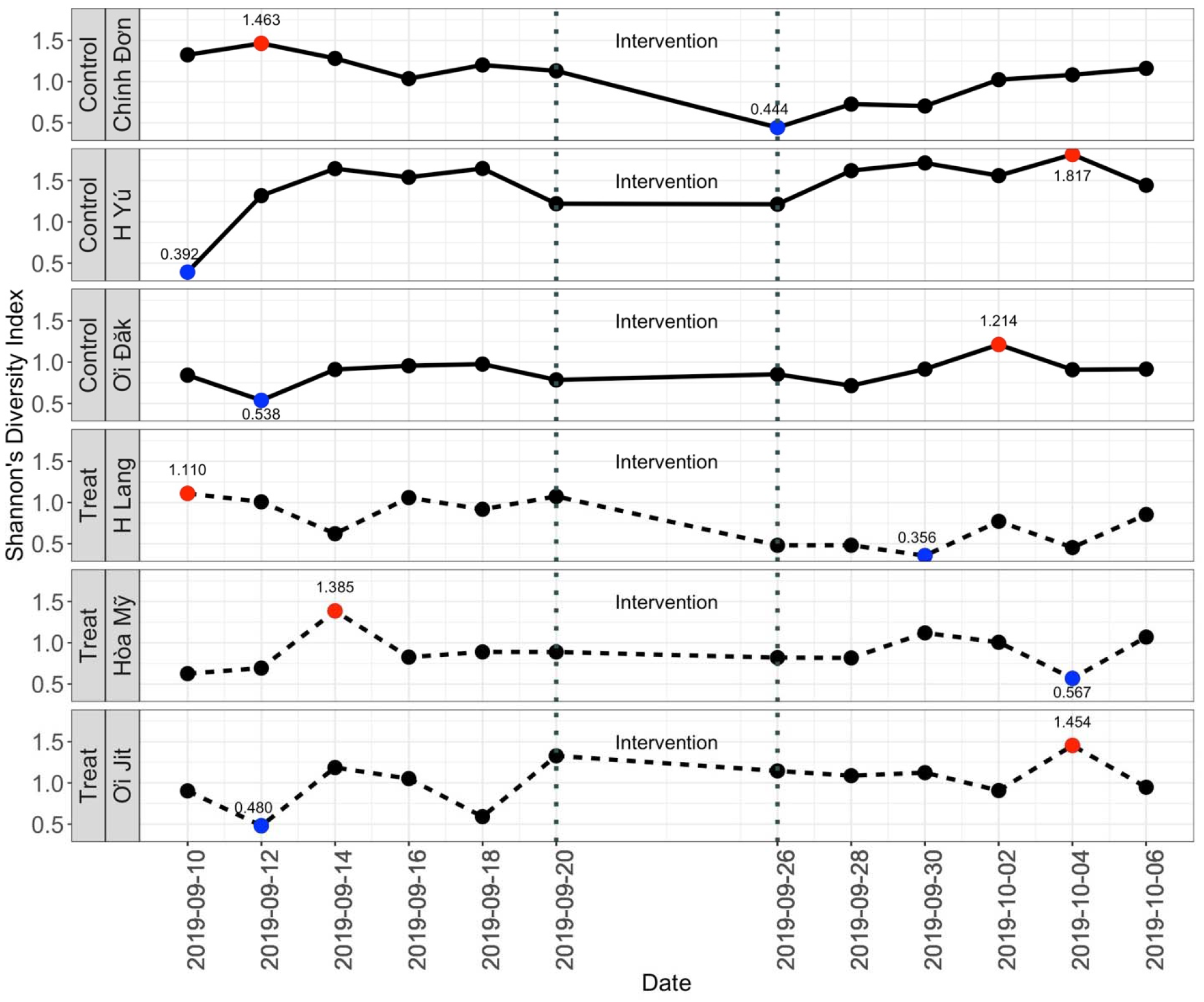
Shannon’s index calculated for each day of mosquito captures Shannon’s index was calculated for each date in each village. Red circles represent the maximum value and blue circles represent the minimum value. Higher values indicate increased levels of diversity.

**Figure 5:**
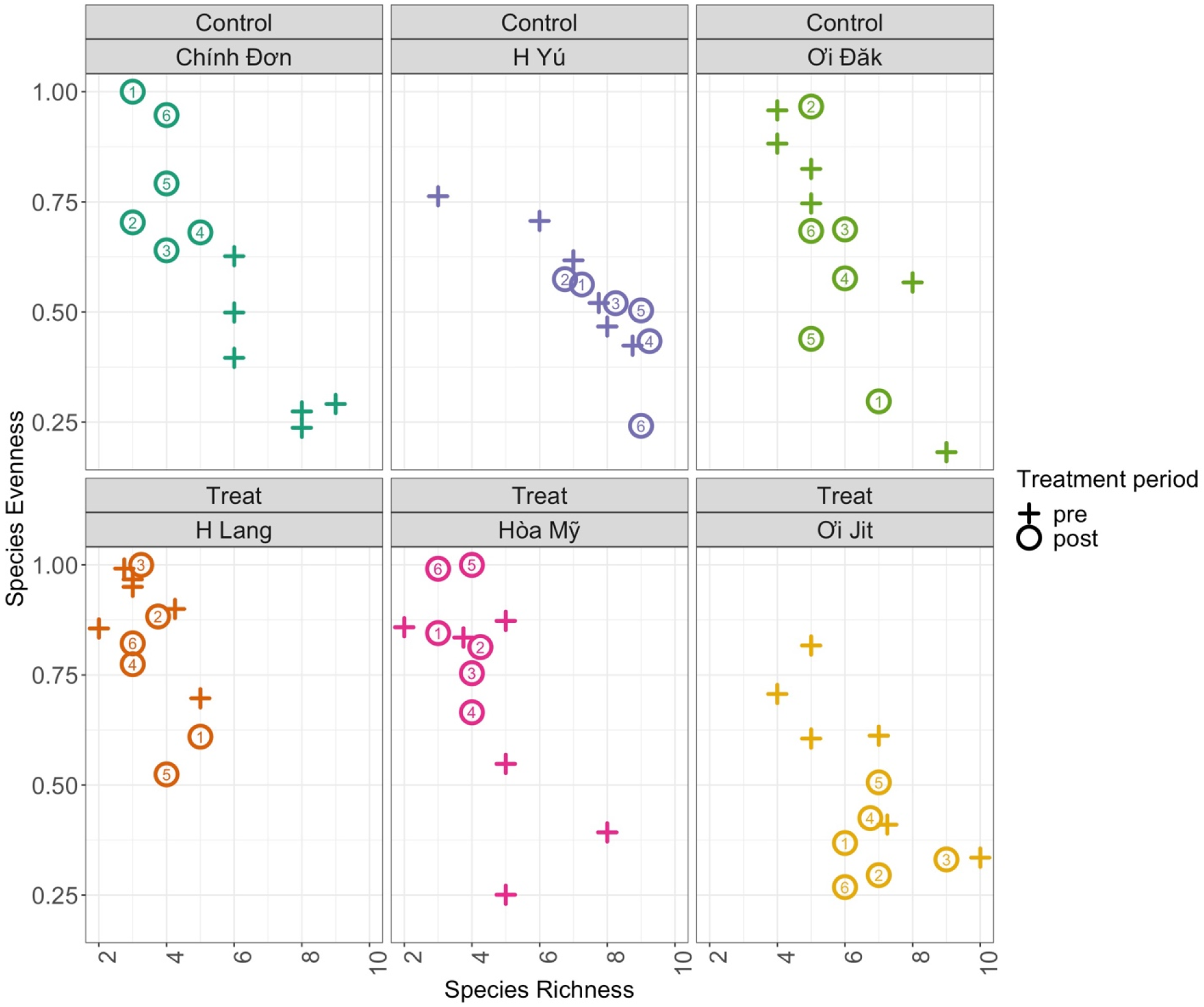
Decomposition of Shannon’s Diversity Index into richness and evenness This figure represents the calculated richness and evenness for each village on each date. Crosses represent the dates before treatment, and circles represent the dates after treatment. Numbers in circles represent the number of days after treatment. Higher values on the y-axis represent increased species diversity. Higher values on the x-axis represent increased species richness.

Of the villages that experienced significant changes in the Shannon diversity index, two showed increased diversity (HY (control), and OJ (treat)) and two showed significantly less diversity (CD (control) and HL (treat)) overall (Figure 3, Table 3). When diversity was stratified by trap-night, there were no apparent trends in daily species diversity. The maximum value for the diversity index occurred in three villages (control village CD and treatment villages HL and HM) during the pre-treatment period, and in three villages (control villages HY and OD, and treatment village OJ) during the post-treatment period. Additionally, in each village site, there was no clear change in trend due to the intervention (Figure 4). Of the three control villages, two (HY and OD) had a local minimum before treatment and one (CD) a local maximum before treatment. Of the treatment villages, two (HM and HL) had a local maximum before treatment and one (OJ) had a local minimum. Though in some villages such as HL there appears to be less diversity after treatment, the change in diversity is not consistent throughout the treatment or control groups.

**Table 3:** *Anopheles* species diversity metrics

| Village Name | Study arm | Shannon's pre-intervention | Shannon's post-intervention | p-value for difference* | Total Species (pre-) | Total Species (post-) |
| --- | --- | --- | --- | --- | --- | --- |
| Chính Đơn (CD) | Control | 1.28 | 1.07 | 0.002 | 13 | 6 |
| H Lang (HL) | Treat | 1.56 | 0.61 | <.0001 | 7 | 7 |
| H Yú (HY) | Control | 1.45 | 1.68 | 0.014 | 11 | 13 |
| Hòa Mỹ (HM) | Treat | 1.19 | 1.10 | 0.352 | 11 | 8 |
| Ơi Đăk (OD) | Control | 1.04 | 1.01 | 0.661 | 11 | 9 |
| Ơi Jit (OJ) | Treat | 1.04 | 1.18 | 0.013 | 11 | 10 |

When analyzing species evenness and richness individually, there was no apparent trend across days post-intervention or across treatment vs. control grouping. Some villages exhibited more species richness during the pre-treatment period and higher species evenness of species evenness during the post-treatment period, however, there does not appear to be a trend that occurred over all villages in the treatment or control groups (Figure 5).

## Discussion

The results of this study did not provide statistically-significant evidence that treating cattle in villages with IVM reduces nightly captures from cattle-baited traps of female *Anopheles* mosquitoes, compared to cattle in villages not treated with IVM. There were no consistent impacts of IVM on the *Anopheles* species diversity; three villages exhibited less diversity while three villages experienced greater diversity after the intervention period. Moreover, large decreases in nightly captures were measured during the study across both intervention and control villages. This lack of differential impacts may be due to combinations of the factors as discussed below.

### The duration of IVM activity in treated cattle may be insufficient for population-level impacts

This could be remedied through the use of long-release IVM treatments such as LongRange,^31^ which have the potential to expose a larger number of mosquitoes to IVM before it is cleared from the cattle.

### There was the potential for crossover in mosquito populations between treated and control villages

The villages in this study population were in some proximity to each other. It is possible that mosquitoes fed on cattle in areas without IVM treatment and were then captured in CBTs in treatment villages. It is also possible that the converse happened; mosquitoes fed on IVM-treated cattle and then moved to control villages. However, this is less likely because the IVM has a higher chance of causing mortality in mosquitoes before they move to control villages as they need to rest to digest the bloodmeal. This scenario is supported by the decrease in mosquitoes captured in both the treatment and control villages during the post- vs. pre-period. During a capture-recapture study of *An. maculatus* in Malaysia, 68% of recaptures were taken within a distance of 0.5 km, however, a flight range of 1.6 km was detected; ^32^ this species is a known vector in adjacent areas of Vietnam.^33^

### Mosquitoes may not have fed on cattle

Many vectors in this population are largely anthropophilic and thus may not readily have fed on the cattle. The most common mosquito population captured was *An. peditaeniatus* which is primarily anthropophilic^17^ and therefore is less likely to be impacted by animal-based interventions. However, CBTs were used for the primary outcome metric, thus it seems unlikely that the lack of impact of IVM was due to mosquitoes selecting hosts other than the cattle.

### Spatial spillover

In addition to vector movement between villages, it is possible that the treated cattle may have impacted vectors outside their assigned study arm. This would be problematic if herd movements coincided with crepuscular feeding, which is common in the GMS.^34^ If the cattle are only grazing outside of their randomized area during the day (when few vectors are active) this would be unlikely to impact the outcomes; however, if cattle are grazing outside of their randomized area during dawn or dusk when there is increased vector biting, this would lead to biased results.^35^ To account for the lack of clear spatial boundaries caused by cattle grazing in different locations, spatial impacts would need to be included in both the analyses and the design of a future study to account for cattle movement and vector migration.^35,36^

### Extensive mosquito trapping may have impacted mosquito populations

The villages in this study ranged in size from approximately 0.0998 km^2^ (OD) to 1.0640 km^2^ (HL). In these small areas, the mosquito collection may have impacted the number of mosquitoes in the population, thus trapping itself may have had a larger impact on nightly mosquito captures than the intervention, as large impacts were shown in earlier studies with African vectors.^37^ Additionally, population periodicity may have played a role in mosquitoes captured. Adult and larval mosquitoes have uncorrelated development stages, therefore it is possible there were insufficient larvae to replenish the adult population during this study experiment.^38^

### Impact of moonlight

Animal baited-traps have been shown to be impacted by ambient moonlight.^39^ The major peak of trapping (Sept 14 coincided with a full moon) and the nadir was associated with a new moon (Sept 28).

Strengths of this study include the fact that it is the first study, to our knowledge, to examine the effect of IVM-treated cattle on wild anopheline populations in a field setting. Though prior work has shown that IVM is an effective mosquitocide in laboratory conditions,^13^ no prior work has shown its effectiveness in a field trial. Limitations of this study include the limited number of villages included in the trial design; however, it was well-powered for primary outcomes. Future studies looking at the effectiveness of IVM on a village level should consider including more villages with expansive spatial buffers. Additionally, in the village HL, coverage was lower than the desired 80%. Future studies should devise plans to treat at least 80% of the cattle in the village in order to best evaluate results. Additionally, future work should prioritize the measurement of the relationship between IVM dose-density over a defined catchment and associated mosquito capture rates. This would be an important index to determine thresholds for the density of IVM-treated cattle needed to impact mosquito captures.

Results from this pilot study did not show evidence for statistically significant differences in total anopheline captures in IVM-treated villages relative to control villages. The marked decrease in captures across all study sites post-intervention limited our ability to quantify changes due to IVM treatments. These changes may be due to natural population fluctuations; spill-over of treated cows between villages; movement of treated vectors; or due to saturation of trapping around the central cattle-baited traps. Future work should prioritize fully spatially-separated clusters (400m minimum); non-static trapping stations for outcomes; and more controlled monitoring of cattle movements. If IVM treatment is to be an effective tool in reducing vector populations, it is vital to understand how to best scale this intervention for population-level epidemiological impacts.

## Data Availability

Data will be made publically available at osf.io

## Acknowledgments

We thank all the dedicated staff members at the Institute of Malariology, Parasitology, and Entomology – Quy Nhon for their partnership in this project.

## Disclosures regarding real or perceived conflicts of interest

JCH is a military service member of the United States government. This work was prepared as part of his official duties. Title 17 U.S.C. 105 provides that ‘copyright protection under this title is not available for any work of the United States Government.’ Title 17 U.S.C. 101 defines a U.S. Government work as work prepared by a military service member or employee of the U.S. Government as part of that person’s official duties. The views expressed in this article reflect the results of research conducted by the authors and do not necessarily reflect the official policy or position of the Department of the Navy, Defense Health Agency, Department of Defense, nor the United States Government.

## Tables and Figures

**Supplemental Table 1.** Sample size analysis.

| Trapping nights,<br>pre/post | Trapping stations<br>per village | Mean<br>captures per<br>trap-night | SD of trapping<br>rates | Study<br>power |
| --- | --- | --- | --- | --- |
| 6/6 | 1 | 35 | 20 | 0.79 |
| 6/6 | 1 | 40 | 20 | 0.84 |
| 7/7 | 1 | 35 | 20 | 0.84 |
| 7/7 | 1 | 40 | 20 | 0.90 |

